# Phenotypic Classification of Multisystem Inflammatory Syndrome in Children: A Latent Class Analysis

**DOI:** 10.1101/2024.06.01.24308325

**Authors:** Kevin C. Ma, Anna R. Yousaf, Allison Miller, Katherine N. Lindsey, Michael J. Wu, Michael Melgar, Ami B. Popovich, Angela P. Campbell, Laura D. Zambrano

## Abstract

**Importance:** Multisystem inflammatory syndrome in children (MIS-C) is an uncommon but severe hyperinflammatory illness occurring 2–6 weeks after SARS-CoV-2 infection. Presentation overlaps with other conditions, and risk factors for severe clinical outcomes differ by patient. Characterizing patterns of MIS-C presentation can guide efforts to reduce misclassification, categorize phenotypes, and identify patients at risk for severe outcomes.

**Objective:** To characterize phenotypic clusters of MIS-C and identify clusters with increased clinical severity.

**Design:** We describe MIS-C phenotypic clusters inferred using latent class analysis (LCA) applied to the largest cohort to date of cases from U.S. national surveillance. Illness onset ranged from February 2020 through December 2022.

**Setting:** National surveillance comprising data from 55 U.S. public health jurisdictions.

**Participants:** We analyzed 9,333 MIS-C cases. Twenty-nine clinical signs and symptoms were selected for clustering after excluding variables with ≥20% missingness and ≤10% or ≥90% prevalence. We excluded 389 cases missing ≥10 variables and conducted multiple imputation on the remaining 8,944 (96%) cases.

**Main Outcomes and Measures:** Differences by cluster in prevalence of each clinical sign and symptom, percentage of cases admitted to the intensive care unit (ICU), length of hospital and ICU stay, mortality, and relative frequency over time.

**Results:** LCA identified three clusters characterized by 1) frequent respiratory findings primarily affecting older children (n = 713; 8.0% of cases; median age: 12.7 years); 2) frequent cardiac complications and shock (n = 3,359; 37.6%; 10.8 years); and 3) remaining cases (n = 4,872; 54.5%; 6.8 years). Mean duration of hospitalization and proportion of cases resulting in ICU admission or death were higher in the respiratory (7.9 days; 49.5%; 4.6%; respectively) and shock/cardiac clusters (8.7 days; 82.3%; 1.0%; respectively) compared with other cases (5.3 days; 33.0%; 0.06%; respectively). The proportion of cases in the respiratory and shock/cardiac clusters decreased after emergence of the Omicron variant in the United States.

**Conclusions and Relevance:** MIS-C cases clustered into three subgroups with distinct clinical phenotypes, illness severity, and distribution over time. Use of clusters in future studies may support efforts to evaluate surveillance case definitions and help identify groups at highest risk for severe outcomes.

**Key points:** *Question:* Can phenotypic clusters of multisystem inflammatory syndrome in children (MIS-C) be identified, and are some clusters associated with increased severity?

*Findings:* We describe clusters inferred using latent class analysis (LCA) on 9,333 MIS-C cases from U.S. national surveillance 2020–2022. LCA identified three clusters characterized by frequent respiratory symptoms, frequent cardiac complications and shock, and remaining clinically milder cases. Mortality and ICU admission were highest in the respiratory and shock/cardiac clusters; prevalence of these two clusters decreased over time.

*Meaning:* MIS-C clusters had distinct presentation, illness severity, and distribution over time, highlighting the importance of recognizing the varied presentation of MIS-C.

## Introduction

Multisystem inflammatory syndrome in children (MIS-C) is an uncommon but severe hyperinflammatory syndrome occurring 2–6 weeks after SARS-CoV-2 infection, postulated to arise because of post-infectious immune dysregulation [1,2]. As of April 29, 2024, there have been 9,684 MIS-C cases and 79 associated deaths in the United States reported to the U.S. Centers for Disease Control and Prevention (CDC) national MIS-C surveillance system, with higher incidence observed among racial and ethnic minorities early in the pandemic [3–5]. Patients with MIS-C commonly present with fever, gastrointestinal symptoms including vomiting, diarrhea, and abdominal pain, cardiac dysfunction, and mucocutaneous symptoms including rash and conjunctivitis [6]. In May 2020, CDC developed a surveillance case definition for MIS-C using the following criteria: patient <21 years with fever and illness requiring hospitalization, involvement of ≥2 organ systems, laboratory evidence of inflammation, laboratory or epidemiologic evidence of current or recent SARS-CoV-2 infection, and no alternative plausible diagnosis [7,8]. This definition was broad to maximize MIS-C case capture. CDC and the Council for State and Territorial Epidemiologists (CSTE) created a new MIS-C surveillance case definition, effective January 2023, that aimed to reduce misclassification and minimize reporting burden [9]. Changes included removal of the respiratory organ involvement criterion to potentially decrease misclassification of severe acute pediatric COVID-19 as MIS-C.

MIS-C case finding potentially captures a range of conditions because presentation shares clinical features with other conditions including acute COVID-19, Kawasaki disease, and toxic shock syndrome, which may complicate timely diagnosis and treatment [6,10,11]. Progression to MIS-C severe outcomes (including the occurrence of shock, intensive care unit (ICU) admission, and death) can also vary substantially, with some signs and symptoms associated with increased severity [12,13]. However, while some individual risk factors have been characterized, understanding of the drivers of severity remains incomplete.

This heterogeneity in presentation and clinical severity suggests the existence of phenotypic classes of MIS-C, as has been documented for other conditions including acute respiratory distress syndrome [14]. Characterizing these classes could improve understanding of MIS-C pathophysiology, guide subgroup analyses in clinical trials of therapeutic options, and support refinement of case definitions and continued epidemiologic surveillance. Previous studies have employed latent class analysis (LCA) and other clustering algorithms to characterize the spectrum of MIS-C presentation [7,10,15]. These analyses have informed MIS-C case definition updates and highlight the potential of applying statistical models to interpret MIS-C heterogeneity [9,16], but have been limited in sample size and geographic representativeness. Additionally, some prior clustering studies have aggregated input clinical data into broad organ system categories, which loses information on both severity of symptoms and the number of symptoms occurring within organ system groups.

The landscape of MIS-C incidence, surveillance, and treatment has changed considerably since SARS-CoV-2 first emerged, highlighting the need for an updated categorization of MIS-C phenotypic clusters. Overall incidence of MIS-C has decreased as new SARS-CoV-2 variants such as Omicron and its descendants have emerged [17–22], but increases in MIS-C cases can still occur following surges in COVID-19 transmission [23,24]. Here, we apply statistical clustering methods on the largest cohort of U.S. MIS-C cases to date to systematically characterize MIS-C phenotypic clusters and investigate their association with clinical severity.

## Methods

### Study design and cohort

CDC started national passive surveillance for MIS-C cases on May 14, 2020. U.S health departments voluntarily report MIS-C cases using a standardized case report form that includes questions on patient demographics, clinical signs and symptoms, treatments, and imaging and laboratory results [25]. We conducted a retrospective analysis of all cases reported to national surveillance as of April 4, 2023 with MIS-C symptom onset on or prior to December 31, 2022 meeting the May 2020 CDC MIS-C surveillance case definition [7,8]. We excluded cases with onset January 1, 2023 onwards that were adjudicated with the 2023 CDC/CSTE case definition [9]. Data from free-text responses from the surveillance case report form were reviewed by clinicians and categorized appropriately. This activity was reviewed by CDC and conducted consistent with applicable federal law and CDC policy (see e.g., 45 C.F.R. part 46.102(l)(2), 21 C.F.R. part 56; 42 U.S.C. §241(d); 5 U.S.C. §552a; 44 U.S.C. §3501 et seq.).

We conducted descriptive analysis of characteristics of patients by SARS-CoV-2 variant predominance periods. Periods were defined using ≥50% SARS-CoV-2 variant proportions from national genomic surveillance [26,27] with an additional two week lag due to presumed minimal delay from SARS-CoV-2 infection to MIS-C onset: pre-Delta (July 9, 2021, and earlier); Delta (July 10–December 31, 2021); Omicron (January 1, 2022, and later).

## Statistical analysis

### Indicator variable selection

We selected clinical signs and symptoms, diagnoses, complications, imaging results, and laboratory testing variables from the case report form for use as binary indicator (i.e., input) variables in LCA (Supplementary File 1). To restrict to the most informative indicators, we removed variables with a high percentage (≥20%) of missing data, high correlation with other indicator variables (as defined by Pearson correlation ≥0.5), and rare (≤10%) or high (≥90%) prevalence. Prevalence was calculated based on non-missing data except for variables derived from echocardiogram results, where prevalence was calculated among all case patients. Missingness for echocardiogram results was likely not at random as selection of which patients received an echocardiogram could depend on clinical judgement of the extent of cardiac involvement. We additionally created composite variables for overlapping or redundant fields, resulting in 29 final variables (Supplementary Methods).

Cases with ≥10 missing variables (n = 389) were excluded and multiple imputation of the 29 clinical variables was conducted on all remaining 8,944 cases (95.8% of all cases). Clinical outcomes of interest (including ICU admission, ICU and hospitalization lengths of stay, and mortality), patient demographics, and comorbidities were not used as indicator variables and did not influence cluster assignment.

### Latent class analysis and interpretation

LCA was run using the *poLCA* R package (number of times to estimate the model set to 20 and maximum iterations set to 5,000 with default parameters otherwise) and LCA combined with variable selection was run using the *LCAvarsel* package [28,29]. The number of clusters was selected after considering multiple factors, including 1) model fit based on the Bayesian Information Criterion and Akaike Information Criterion, 2) assessment of class distinctiveness using entropy and principal component analysis (PCA), a variable reduction approach intended to simplify complex datasets into a smaller number of variables (i.e., principal components) that contribute the most variance [30], 3) comparability with results from integrating LCA with variable selection, 4) evaluation of the consistency of clustering solutions by running LCA on random subsamples of our dataset, and 5) clinical interpretability (Supplementary Methods).

### Cluster interpretation

We interpreted clusters based on distinct clinical signs, symptoms, complications, and laboratory testing results. We performed descriptive analyses of differences between clusters by patient demographics, comorbidities, and clinical outcomes (ICU admission, ICU and hospitalization lengths of stay, and mortality). Statistical significance of global differences across clusters was assessed using the Chi-square test for categorical variables and Kruskal-Wallis test for continuous variables. *P-*values less than 0.05 were interpreted as evidence that distributions were significantly different between clusters. Changes in the prevalence of clusters over time were described across SARS-CoV-2 predominant variant periods.

To estimate differences in case ascertainment between the 2023 CSTE/CDC surveillance case definition and the 2020 case definition, we calculated the percentage of MIS-C cases in each cluster that would also meet the 2023 CSTE/CDC surveillance case definition, excluding cases without a reported quantitative CRP measurement.

R software (version 4.1.3; R Foundation) was used to conduct all analyses.

## Results

We analyzed 8,944 MIS-C cases meeting inclusion criteria and reported to CDC using the 2020 CDC MIS-C surveillance case definition. Symptom onset ranged from February 19, 2020, through December 31, 2022. Case patients had a median age of 8.7 years (IQR 5.0–13.0 years) and the majority were male (n = 5,407; 60.5%) (Table 1). Pre-existing conditions were reported for 2,210 (24.7%) patients, with the most common conditions being obesity (n = 1,287; 14.4%), chronic lung disease (n = 577; 6.5%), and congenital malformations excluding congenital heart disease (n = 362; 4.0%). Approximately half of reported MIS-C patients (n = 4,727; 52.9%) were admitted to the ICU and 70 (0.8%) died. Among the included MIS-C cases, the percentages reported during the pre-Delta, Delta, and Omicron variant predominant periods were 58.0%, 25.9%, and 16.1%, respectively. Among case patients in each variant period, the percentages admitted to the ICU were 57.4%, 50.3%, and 40.5%, respectively, and median age decreased from 9.0 years (IQR 5.1–13.0) during the pre-Delta period to 7.2 years (IQR 3.9–11) during the Omicron period (*P*<.001).

**Table 1.**
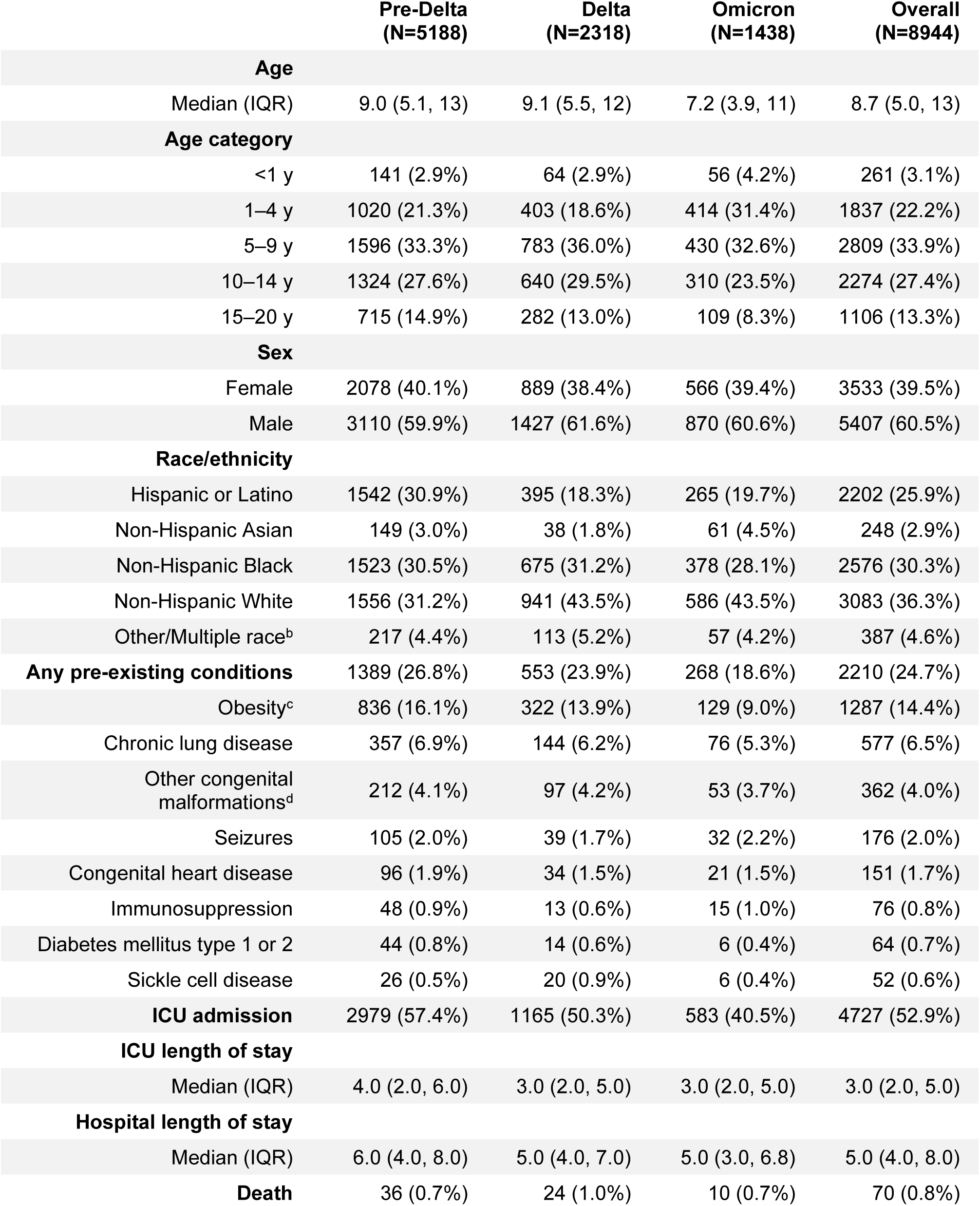
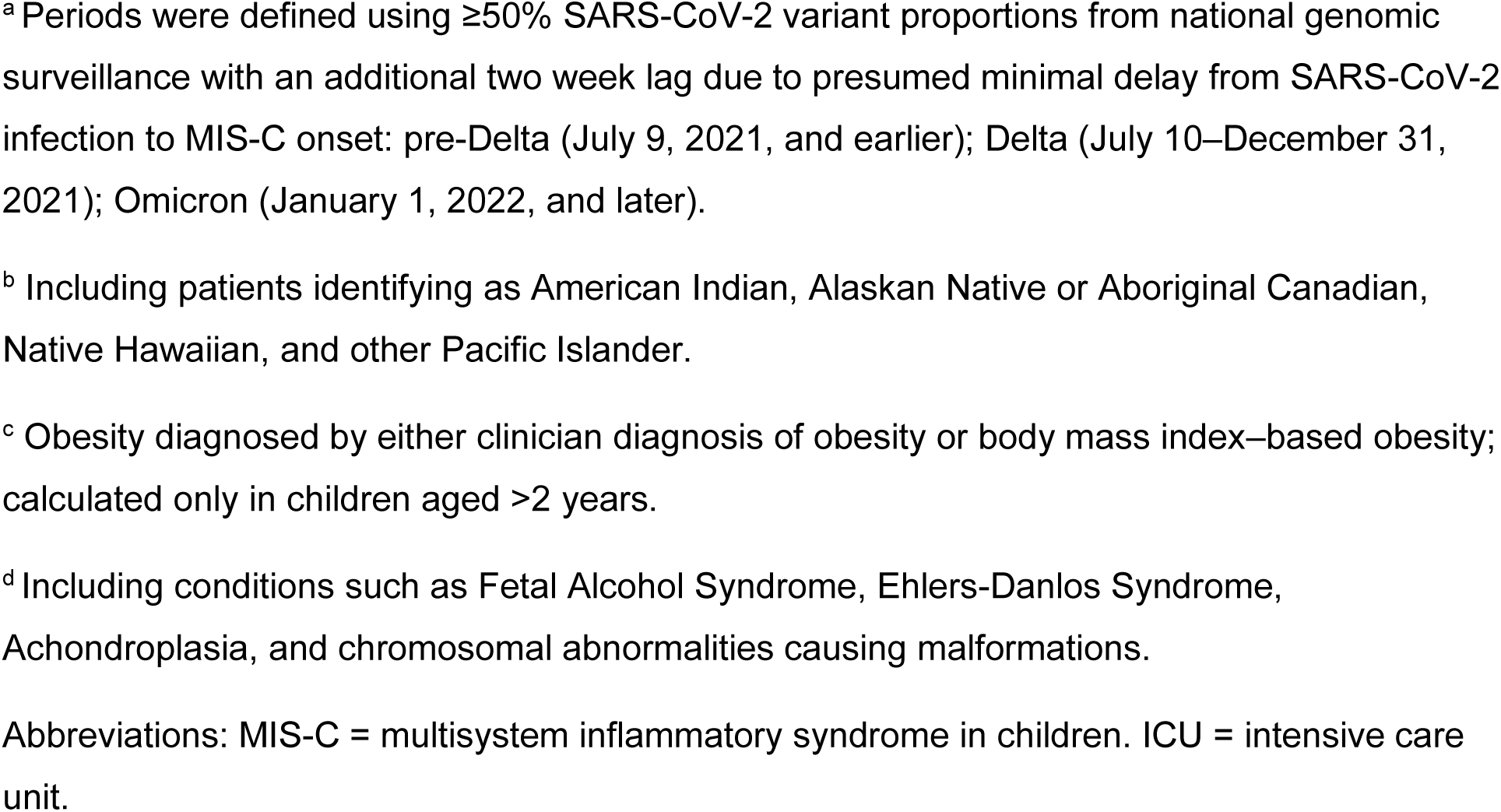
Characteristics of MIS-C patients by predominant circulating SARS-CoV-2 variant^a^ — United States, February 2020–December 2022.

|  | **Pre-Delta**<br>**(N=5188)** | **Delta**<br>**(N=2318)** | **Omicron**<br>**(N=1438)** | **Overall**<br>**(N=8944)** |
| --- | --- | --- | --- | --- |
| **Age** |  |  |  |  |
| Median (IQR) | 9.0 (5.1, 13) | 9.1 (5.5, 12) | 7.2 (3.9, 11) | 8.7 (5.0, 13) |
| **Age category** |  |  |  |  |
| <1 y | 141 (2.9%) | 64 (2.9%) | 56 (4.2%) | 261 (3.1%) |
| 1–4 y | 1020 (21.3%) | 403 (18.6%) | 414 (31.4%) | 1837 (22.2%) |
| 5–9 y | 1596 (33.3%) | 783 (36.0%) | 430 (32.6%) | 2809 (33.9%) |
| 10–14 y | 1324 (27.6%) | 640 (29.5%) | 310 (23.5%) | 2274 (27.4%) |
| 15–20 y | 715 (14.9%) | 282 (13.0%) | 109 (8.3%) | 1106 (13.3%) |
| **Sex** |  |  |  |  |
| Female | 2078 (40.1%) | 889 (38.4%) | 566 (39.4%) | 3533 (39.5%) |
| Male | 3110 (59.9%) | 1427 (61.6%) | 870 (60.6%) | 5407 (60.5%) |
| **Race/ethnicity** |  |  |  |  |
| Hispanic or Latino | 1542 (30.9%) | 395 (18.3%) | 265 (19.7%) | 2202 (25.9%) |
| Non-Hispanic Asian | 149 (3.0%) | 38 (1.8%) | 61 (4.5%) | 248 (2.9%) |
| Non-Hispanic Black | 1523 (30.5%) | 675 (31.2%) | 378 (28.1%) | 2576 (30.3%) |
| Non-Hispanic White | 1556 (31.2%) | 941 (43.5%) | 586 (43.5%) | 3083 (36.3%) |
| Other/Multiple race<sup>b</sup> | 217 (4.4%) | 113 (5.2%) | 57 (4.2%) | 387 (4.6%) |
| **Any pre-existing conditions** | 1389 (26.8%) | 553 (23.9%) | 268 (18.6%) | 2210 (24.7%) |
| Obesity<sup>c</sup> | 836 (16.1%) | 322 (13.9%) | 129 (9.0%) | 1287 (14.4%) |
| Chronic lung disease | 357 (6.9%) | 144 (6.2%) | 76 (5.3%) | 577 (6.5%) |
| Other congenital malformations<sup>d</sup> | 212 (4.1%) | 97 (4.2%) | 53 (3.7%) | 362 (4.0%) |
| Seizures | 105 (2.0%) | 39 (1.7%) | 32 (2.2%) | 176 (2.0%) |
| Congenital heart disease | 96 (1.9%) | 34 (1.5%) | 21 (1.5%) | 151 (1.7%) |
| Immunosuppression | 48 (0.9%) | 13 (0.6%) | 15 (1.0%) | 76 (0.8%) |
| Diabetes mellitus type 1 or 2 | 44 (0.8%) | 14 (0.6%) | 6 (0.4%) | 64 (0.7%) |
| Sickle cell disease | 26 (0.5%) | 20 (0.9%) | 6 (0.4%) | 52 (0.6%) |
| **ICU admission** | 2979 (57.4%) | 1165 (50.3%) | 583 (40.5%) | 4727 (52.9%) |
| **ICU length of stay** |  |  |  |  |
| Median (IQR) | 4.0 (2.0, 6.0) | 3.0 (2.0, 5.0) | 3.0 (2.0, 5.0) | 3.0 (2.0, 5.0) |
| **Hospital length of stay** |  |  |  |  |
| Median (IQR) | 6.0 (4.0, 8.0) | 5.0 (4.0, 7.0) | 5.0 (3.0, 6.8) | 5.0 (4.0, 8.0) |
| **Death** | 36 (0.7%) | 24 (1.0%) | 10 (0.7%) | 70 (0.8%) |

<sup>a</sup> Periods were defined using $\geq 50\%$ SARS-CoV-2 variant proportions from national genomic surveillance with an additional two week lag due to presumed minimal delay from SARS-CoV-2 infection to MIS-C onset: pre-Delta (July 9, 2021, and earlier); Delta (July 10–December 31, 2021); Omicron (January 1, 2022, and later).
<sup>b</sup> Including patients identifying as American Indian, Alaskan Native or Aboriginal Canadian, Native Hawaiian, and other Pacific Islander.
<sup>c</sup> Obesity diagnosed by either clinician diagnosis of obesity or body mass index–based obesity; calculated only in children aged $>2$ years.
<sup>d</sup> Including conditions such as Fetal Alcohol Syndrome, Ehlers-Danlos Syndrome, Achondroplasia, and chromosomal abnormalities causing malformations.
Abbreviations: MIS-C = multisystem inflammatory syndrome in children. ICU = intensive care unit.

We conducted latent class analysis (LCA) on the 8,944 MIS-C cases using 29 clinical variables and identified three clusters as a suitable fit after considering multiple factors, including information criteria, cluster distinctiveness, and clinical interpretability (Supplementary Methods). The number and percentages of patients within clusters one, two, and three were 713 (8.0%), 3,359 (37.6%), and 4,872 (54.5%), respectively. We conducted sensitivity analyses on cluster robustness, finding that identified clusters were (1) consistent when subsampling the dataset (Supplementary Figure 3) and (2) similar to clusters obtained from integrating LCA with variable selection (Supplementary Methods).

Case patients in cluster one, termed the respiratory cluster, were characterized by pronounced respiratory organ system involvement (Figure 1). Patients in the respiratory cluster had the highest prevalence of cough, shortness of breath, pneumonia, chest pain/tightness, and acute respiratory distress syndrome (ARDS), and lower prevalence of mucocutaneous symptoms; differences in the prevalence of these signs, symptoms, and diagnoses across the three clusters were statistically significant (*P*<.001 for all characteristics; Figure 1, Supplementary Table 2). We also evaluated differences in demographics and other variables not used in the LCA clustering model. Patients in this cluster tended to be older (median age 13 years; IQR 6– 17 years), and the percentage of patients with one or more comorbidity was 41.2% compared with 31.4% in cluster two and 18.0% in cluster three (*P*<.001). Frequent comorbidities in cluster one patients included obesity (25.8%), non-cardiac congenital malformations (10.0%), and chronic lung disease (9.6%). Percentages of positive SARS-CoV-2 antigen or RT-PCR positive test results and preceding COVID-19–like illness were highest in cluster one (*P*<.001; Supplementary Table 2).

**Figure 1.**
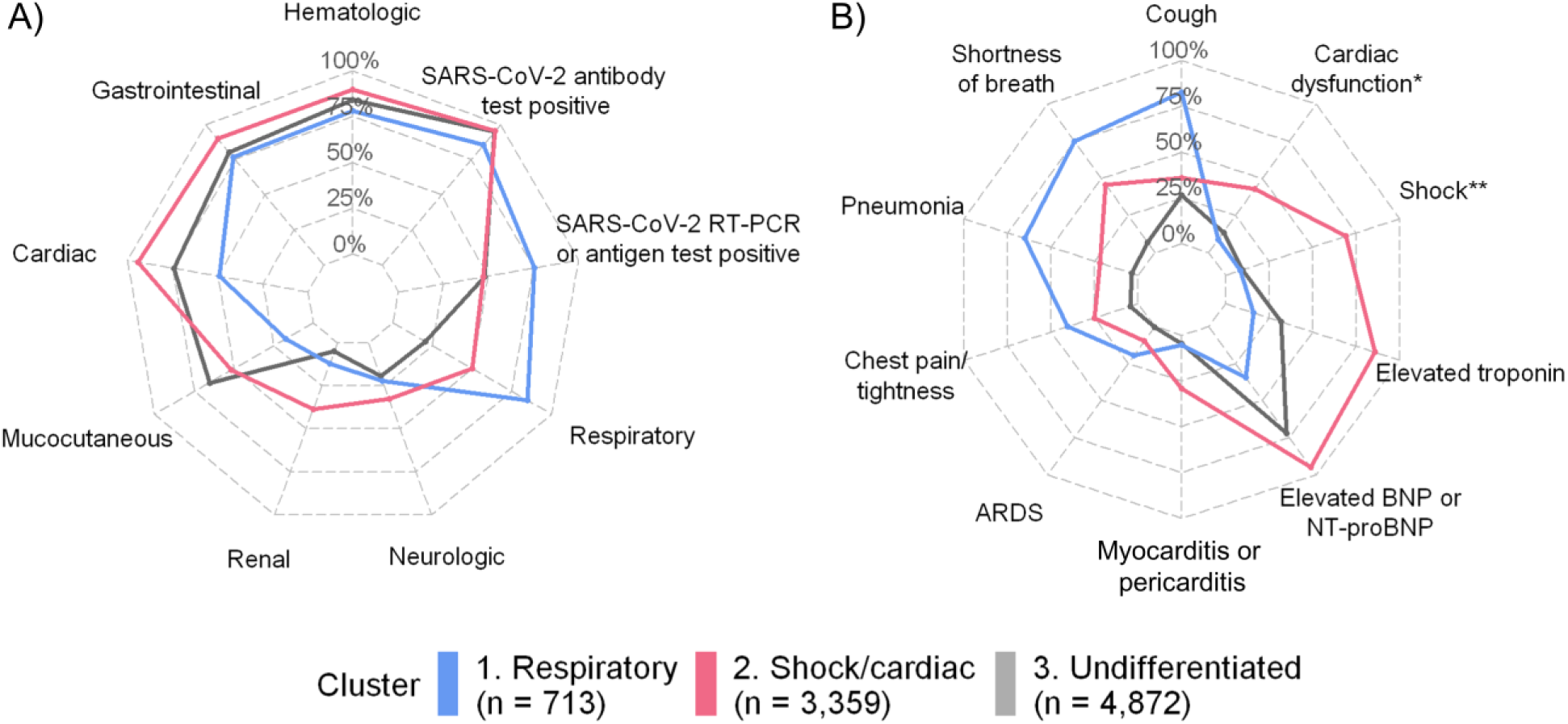
Prevalence of (a) SARS-CoV-2 test results and organ system involvement and (b) common respiratory and cardiac signs, symptoms, diagnoses, or laboratory values by latent class analysis-inferred clusters. Prevalence was calculated based on non-missing data except for cardiac dysfunction, where prevalence was calculated based on all patients in a cluster, as missingness of echocardiogram results was likely not at random. Sample sizes in cluster labels indicate total numbers of patients within each cluster. *Cardiac dysfunction as assessed by left or right ventricular dysfunction on echocardiogram. **Shock indicated in a clinical note or receipt of vasopressors. Abbreviations: ARDS = acute respiratory distress syndrome. BNP = B-type natriuretic peptide. NT-proBNP = N-terminal prohormone of brain natriuretic peptide.

We designated cluster two as the shock/cardiac cluster because patients in this cluster had the highest prevalence of shock (as indicated in a clinical note or receipt of vasopressors; 69.0%) and reported cardiac involvement, comprising any of the occurrence of shock, elevated troponin (dichotomized yes/no), elevated B-type natriuretic peptide (BNP; dichotomized yes/no), abnormal echocardiogram results, or arrhythmia (94.4%) (Figure 1, Supplementary Table 2). Cluster two patients also had the highest prevalence of other cardiac complications such as myocarditis/pericarditis and cardiac dysfunction on echocardiogram (*P*<.001 for both characteristics). Among patients with available data, distributions of BNP (median = 965.0 pg/mL), troponin (median = 0.21 ng/mL), and C-reactive protein (median = 210 mg/L) were elevated relative to other clusters (*P*<.001 for all characteristics; Figure 2). Patients in this cluster also had the largest number of organ systems involved (median of five systems; IQR = 4–5), the highest prevalence of hematologic, gastrointestinal, renal, and neurologic involvement (Figure 1), and slightly younger age (median age 11 years; IQR 8–14 years) compared with respiratory cluster patients (median age 13 years; IQR 6–17 years) (*P*<.001).

**Figure 2.**
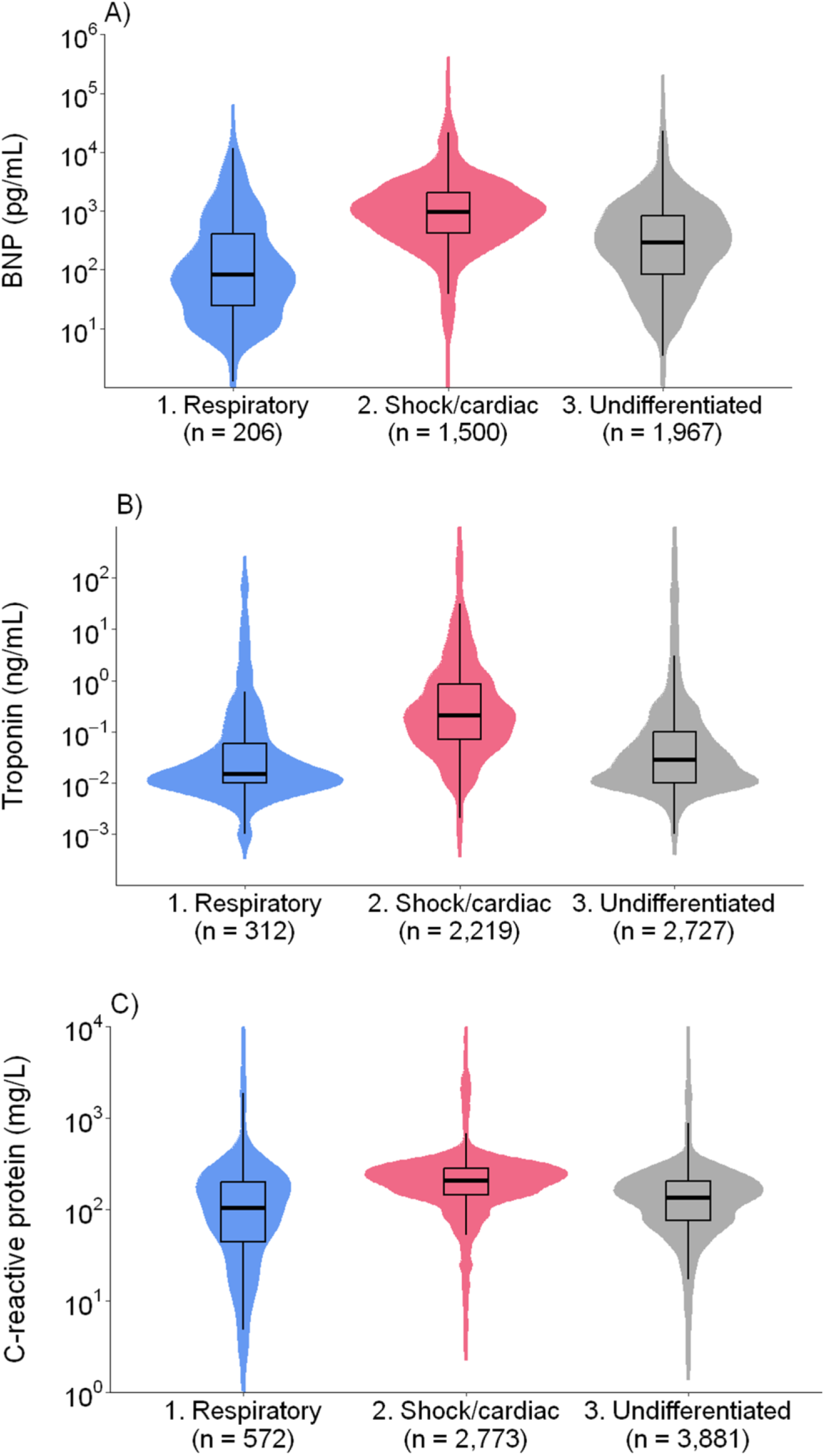
Distributions of levels of (a) B-type natriuretic peptide (BNP), (b) troponin, and (c) C-reactive protein by latent class analysis-inferred clusters. Distributions are scaled to have the same area. Sample sizes indicate numbers of patients within each cluster with non-missing data. Global differences across clusters were statistically significant (*P*<.001) as assessed using the Kruskal-Wallis test for continuous variables for all three clinical biomarkers.

Cluster three comprised the remaining MIS-C case patients who tended to be the youngest (median age 7 years; IQR 4–10 years) with lower numbers of comorbidities (82.0% of patients reported no comorbidities) (Supplementary Table 2). Organ system involvement in this cluster overlapped with the first two clusters, and we designated this the “undifferentiated” cluster (Figure 1). Cluster three patients had slightly higher prevalence of any mucocutaneous symptoms (65.5%) compared with cluster two patients (Figure 1), and 3.4% of patients met clinical criteria for Kawasaki disease (presence of fever, rash, lesions, cervical lymphadenopathy, and conjunctival injection) versus 0% in cluster one and 2.0% in cluster two (*P*<.001).

Morbidity and mortality from MIS-C were concentrated in the respiratory and shock/cardiac clusters. The percentage of MIS-C cases admitted to the ICU was highest for the shock/cardiac cluster (82.3%) followed by the respiratory (49.5%) and undifferentiated clusters (33.0%) (*P*<.001; Figure 3). Similarly, 20.4% of hospitalizations and 19.2% of ICU stays in the respiratory cluster lasted ten or more days compared with 23.0% and 7.7%, respectively, in the cardiac cluster and 6.6% and 1.6%, respectively, in the undifferentiated cluster (Figure 3). The crude case fatality ratios were higher in the respiratory (4.6%) and shock/cardiac clusters (1.0%) compared with the undifferentiated cluster (0.1%; *P*<.001) (Figure 3), and most decedent cases (67/70) belonged to these first two clusters.

**Figure 3.**
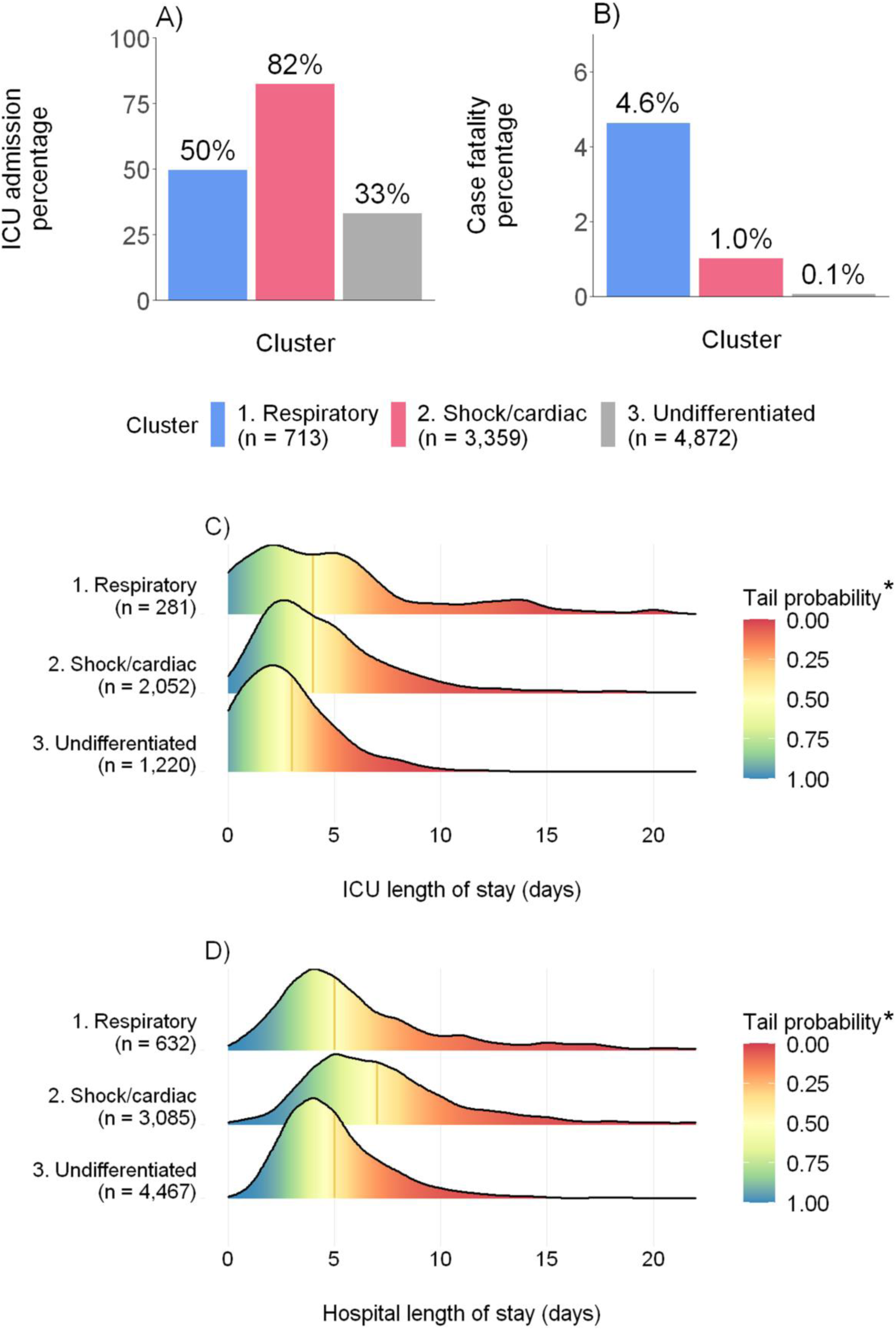
(a) ICU admission percentages, (b) case fatality percentages, and lengths of stay in the (c) ICU and (d) hospital by latent class analysis-inferred cluster. Vertical orange line represents median length of stay in (c) and (d). Sample sizes indicate numbers of patients from each cluster included in the analysis after removing patients with missing data. * Tail probability is defined as the percentage of patients who were in the ICU (c) or hospitalized (d) beyond a certain length of time.

We applied the clusters derived from LCA to characterize the changing epidemiology of MIS-C over time. The combined percentage of MIS-C cases classified in the respiratory and shock/cardiac clusters was 53.0% in May 2020 and remained approximately stable (range = 45.2% to 59.2%) until the end of 2021. This combined percentage began to decrease substantially after emergence of the Omicron variant (Figure 4) and reached 33.2% in October 2022; total counts of monthly MIS-C cases also decreased during this time.

**Figure 4.**
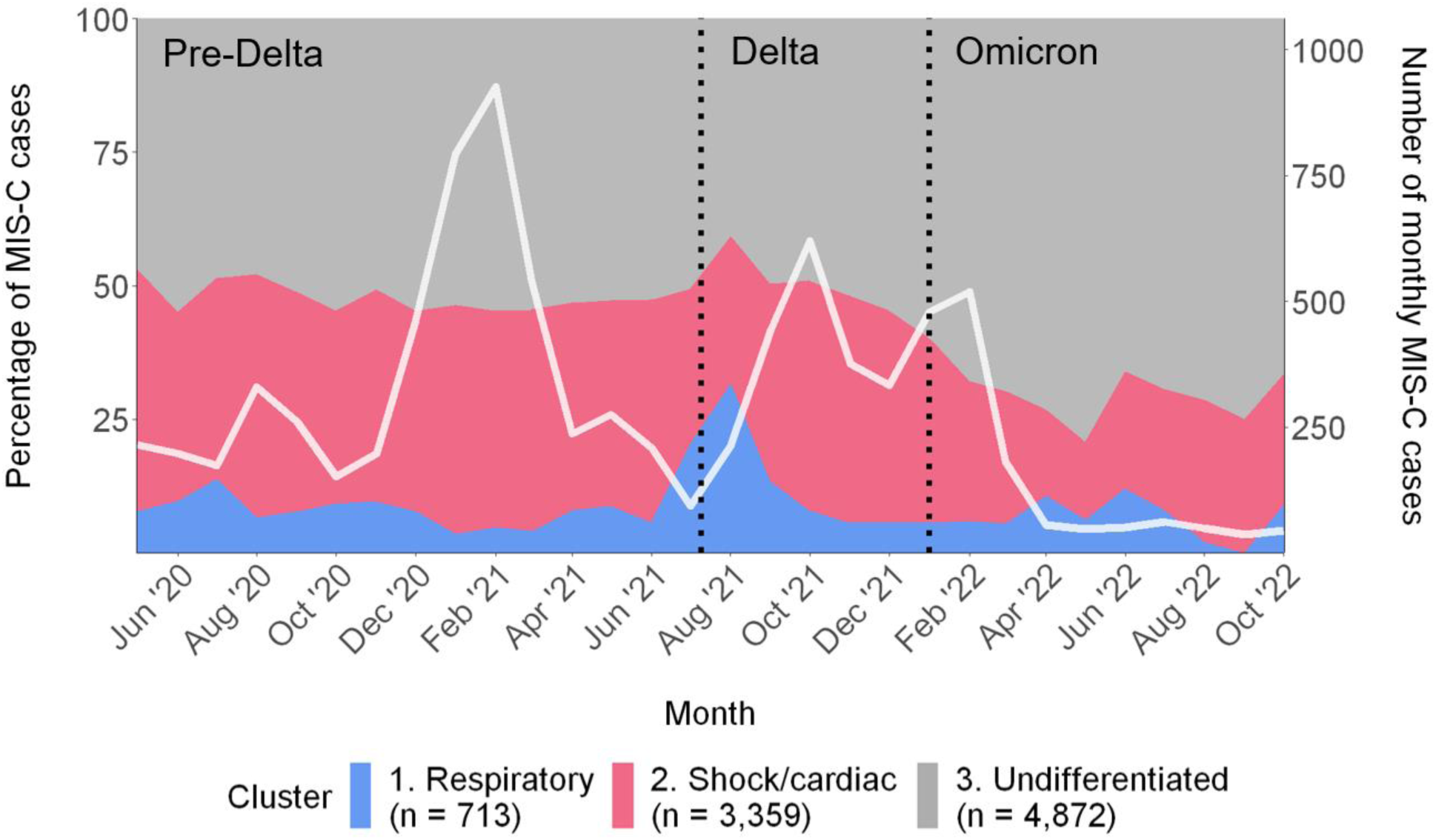
Monthly percentages of MIS-C cases by latent class analysis-inferred cluster (primary y-axis) and number of total MIS-C cases (secondary y-axis) — United States, May 2020–October 2022. Variant periods were defined using ≥50% SARS-CoV-2 variant proportions from national genomic surveillance with an additional two week lag due to presumed minimal delay from SARS-CoV-2 infection to MIS-C onset: pre-Delta (July 9, 2021, and earlier); Delta (July 10–December 31, 2021); Omicron (January 1, 2022, and later) [26,27]. Months in which the total number of cases was less than 30 (April 2020 and prior; November 2022 and onwards) were suppressed per National Center for Health Statistics standards [34].

To evaluate differences in case ascertainment between the 2020 CDC MIS-C surveillance case definition and the 2023 CSTE/CDC case definition, we calculated the percentage of MIS-C cases in each cluster that would meet the 2023 definition. The percentage of case patients meeting the 2023 definition was highest for the shock/cardiac cluster (90.7%) followed by the undifferentiated cluster (77.9%); the percentage in the respiratory cluster was substantially lower (45.8%) (*P<*.001). After restricting to only patients meeting the 2023 case definition, the case fatality ratio among patients in the respiratory (4.9%) and shock/cardiac clusters (1.0%) remained elevated compared to patients in the undifferentiated cluster (0%; *P*<.001).

## Discussion

In this analysis, statistical clustering of MIS-C case patients from U.S. national surveillance with symptom onset February 2020–December 2022 identified three clusters that differed in presentation, clinical outcomes, temporal distribution, and alignment with the 2023 U.S. MIS-C surveillance case definition. Patients in the respiratory cluster comprised older children with comorbidities and a higher prevalence of COVID-19–like illness preceding MIS-C and positive antigen/RT-PCR results. Patients in the shock/cardiac cluster tended to have the highest number of additional organ systems involved. While patients in the undifferentiated cluster had generally less defined organ system involvement, they also had lower clinical severity compared with the other two clusters.

The identification of clusters of MIS-C informs our understanding of heterogeneity in severity and changing epidemiology over time. Previous studies have identified individual risk factors for severity, including shortness of breath, abdominal pain, older age, and increased levels of cardiac dysfunction-associated biomarkers [12]. Our findings support and synthesize these results by demonstrating that clinical severity and mortality is concentrated among two distinct classes of MIS-C patients, defined by predominance of respiratory and shock/cardiac signs and symptoms, respectively. Additionally, decreases in the severity of MIS-C have been described over time [18,19,31,32], and we observed a concurrent decrease in respiratory and shock/cardiac cluster prevalence following emergence of the Omicron variant.

Our results also provide insight into categories of MIS-C patients initially reported using the 2020 CDC surveillance case definition that do or do not also meet criteria for the 2023 CSTE/CDC surveillance case definition [7,9]. The respiratory organ involvement criterion was removed from the 2023 definition in an attempt to decrease misclassification of severe acute COVID-19 as MIS-C. Proportions of discordant patients between the 2020 and 2023 surveillance case definitions were highest for the respiratory cluster and lowest for the shock/cardiac cluster, indicating that the new case definition likely retains sensitivity and specificity for identifying severe MIS-C while potentially reducing misclassification from inclusion of severe acute pediatric COVID-19.

These findings align with and expand upon previous statistical clustering analyses of MIS-C cases. Godfred-Cato et al. identified three clusters among 570 MIS-C cases corresponding to “typical” MIS-C, MIS-C overlapping acute COVID-19, and MIS-C overlapping Kawasaki disease [7]. Geva et al. applied the partitioning around medoids algorithm on 1,526 cases of acute COVID-19 and MIS-C, identifying a “typical” MIS-C cluster with high cardiovascular involvement and hyperinflammation, an acute COVID-19 cluster overlapping MIS-C, and a younger and less critically ill cluster [10]. Rao analyzed 1,139 children using LCA and identified classes corresponding to mild, moderate, and severe presentations [15]. We have identified an MIS-C cluster with high respiratory involvement qualitatively similar to prior studies but with much lower relative frequency compared with corresponding clusters identified in Godfred-Cato et al. and Geva et al. We demonstrated that this relatively rare cluster was associated with increased clinical severity; because this analysis is the largest statistical clustering analysis of MIS-C cases to date, it may have been better powered to identify clusters with lower prevalence and characterize their association with mortality, a relatively rare outcome of MIS-C.

The findings in this report are subject to several limitations. First, because LCA is inherently an exploratory tool, these results are not intended to be used prognostically or for clinical decision-making. Second, we note that the cluster labels we have used here (e.g., respiratory and shock/cardiac clusters) are simplified descriptors; all clusters include some degree of respiratory or cardiac symptoms and cases within clusters are not homogeneous. Third, we opted to use the full range of clinical data available from national surveillance as input variables for LCA, but these correlated data present challenges for LCA, which we tried to address through multiple approaches (Supplementary Methods). Finally, we note that these MIS-C cases were reported using the 2020 CDC MIS-C surveillance case definition and clusters inferred here may not be generalizable to other countries that use different case definitions, or patients reported to CDC in 2023 or later using the 2023 CDC/CSTE case definition [2].

MIS-C remains a public health concern that is likely to accompany surges in SARS-CoV-2 activity, as highlighted by relative increases in case counts during late 2023 compared with prior months [23,24,33]. In this analysis, MIS-C cases reported to CDC clustered into three groups with distinct symptoms and severity, highlighting the importance of recognizing the varied presentation of MIS-C. With additional validation, use of MIS-C phenotypic clusters in public health and clinical settings may be helpful in refining surveillance case definitions, contributing to our understanding of MIS-C pathophysiology, and assisting with recognizing the varied clinical presentations of MIS-C.

## Supporting information

Supplemental Methods and Results

Case report form

## Data Availability

Aggregated data on MIS-C cases reported to U.S. national surveillance are available at: https://covid.cdc.gov/covid-data-tracker/#mis-national-surveillance

## Acknowledgements

Jefferson Jones and Jennifer DeCuir for feedback on clinical interpretation of data; Gordana Derado and Supraja Malladi for statistical support; Joe Abrams, Shana Godfred-Cato, Rebecca Free, and Regina Simeone for project feedback and support on MIS-C national surveillance. We also thank all local, state, and territorial health departments that contributed MIS-C reports to this investigation.

