## Supplemental Methods and Results for "Phenotypic Classification of Multisystem Inflammatory Syndrome in Children: A Latent Class Analysis"

**Supplementary Materials for "Phenotypic Classification of Multisystem Inflammatory Syndrome in Children: A Latent Class Analysis"**

Supplementary Methods……………………………………………………………………….………..2

Supplementary Tables and Figures…………………………………………………………………6

- Supplementary Table 1. Model fit metrics (degrees of freedom, number of parameters, entropy, information metrics, log-likelihood) versus number of clusters………………….6
- Supplementary Table 2. Distribution of demographics, underlying health conditions, clinical signs, symptoms, complications and laboratory testing results, and clinical outcomes by latent class analysis-inferred clusters………………………………………….7

### Supplementary Methods

*Composite variables*

We created composite variables combining identical symptoms present in more than one section of the May 2021 MIS-C case report form. The clinical sign or symptom was considered present if present in any of the aggregated variables:

- Shock: shock during present illness (3.4.1.1), shock as a complication (4.11)
- Pneumonia: pneumonia during present illness (3.4.3.4), pneumonia as a complication (4.8), pneumonia from chest imaging (6.7.2)
- Acute respiratory distress syndrome (ARDS): ARDS during present illness (3.4.3.5), ARDS as a complication (4.7)
- Meningitis or encephalitis: meningitis during present illness (3.4.7.5), encephalitis or aseptic meningitis as a complication (4.10)
- SARS-CoV-2 PCR testing results: inclusion criteria (1.6.1), SARS-CoV-2 testing section (6.8)
- SARS-CoV-2 antibody testing results: inclusion criteria (1.6.2), SARS-CoV-2 testing section (6.10, 6.11, 6.12)
- SARS-CoV-2 antigen testing results: inclusion criteria (1.6.3), SARS-CoV-2 testing section (6.9)

*Indicator variables and excluded variables list*

We removed variables with a high percentage (≥20%) of missing data, high correlation with other indicator variables (as defined by Pearson correlation ≥0.5), and rare (≤10%) or high (≥90%) prevalence. The final list of variables used in the LCA model fitting process were:

- Shock
- Elevated troponin
- Elevated BNP or NT-proBNP
- Arrythmia
- Myocarditis
- Acute kidney injury
- Cough
- Shortness of breath

- Chest pain/tightness
- Pneumonia
- Acute respiratory distress syndrome
- Elevated D-dimers
- Thrombophilia
- Thrombocytopenia
- Abdominal pain
- Vomiting
- Diarrhea
- Elevated bilirubin
- Elevated liver enzymes
- Rash
- Mucocutaneous lesions
- Headache
- Altered mental state
- Syncope/near syncope
- Neck pain
- Myalgia
- Conjunctival injection
- Periorbital edema
- Cervical lymphadenopathy >1.5 cm diameter

The following variables were removed after meeting exclusion criteria:

- High missingness rates: ventricular arrythmia, supraventricular arrythmia, other arrythmia
- Low prevalence: stroke, liver failure, pulmonary embolism, pericarditis, meningitis or encephalitis, encephalopathy, renal failure, congestive heart failure
- High prevalence: fever
- High correlation: hypotension (highly correlated with shock)

*Cluster solution selection for latent class analysis*

We opted to use the full range of clinical signs, symptoms, complications and laboratory testing results available from the MIS-C national surveillance case reporting form as input variables for LCA without collapsing data into broad organ system categories, as has been done in previous LCA analyses. Collapsing multiple symptoms into one category loses information on both severity (e.g., a patient reporting cough is treated equally as a patient experiencing acute respiratory distress syndrome) and the number of symptoms occurring within organ system groups. Additionally, organ system groupings are not distinct boundaries, and symptoms can overlap multiple organ systems. However, conducting LCA on all available symptom data presents challenges, including possible violation of the conditional independence assumption and the identification of spurious latent classes. We therefore used multiple approaches to select the number of clusters and assess robustness of our modeling approach:

1) Information criteria: identifying the number of clusters corresponding to a minimal information criterion is often used to identify best fit LCA models [1]. We observed that with the size of the dataset and number of indicator variables, the Bayesian Information Criterion (BIC) continued to decrease for each additional class added at least up until ten clusters. Instead of a local minimum, we instead looked for an inflection point in the BIC curve (Supplementary Figure 1) as has been suggested in the literature [1]; we observed that the decrease in BIC began to diminish beginning with clusters of size three to five. Similar behavior was observed for other information criteria metrics, including the sample-size adjusted BIC, Akaike Information Criterion (AIC), and consistent AIC (CAIC) (Supplementary Table 1).
2) Cluster distinctiveness: we calculated relative entropy, which is a measure of cluster distinctiveness ranging from 0 to 1 with higher values indicating greater separation between clusters [1]. We found that a solution with three clusters had the highest entropy (Supplementary Table 1; Supplementary Figure 1). We also used principal component analysis, a variable reduction approach intended to simplify complex datasets into a smaller number of variables (i.e., principal components) that contribute the most variance [2]. PCA indicated visual separation in a plot of MIS-C cases along principal components one (x-axis) and two (y-axis) between a three-cluster solution inferred using LCA (Supplementary Figure 2). Principal components one and two explain only 17% of the total variance, and therefore are a limited representation of the full variation in MIS-C clinical phenotypes.

3) Variable selection: we conducted LCA combined with variable selection to characterize the effect of reducing the number of indicator variables on model fit [3]. After running backwards selection with variable swapping via the *LCAvarsel* R package, 12 indicator variables remained. BIC indicated a solution with three or four clusters had the best fit and entropy was again highest for the three-cluster solution, supporting results from LCA model inference conducted without variable selection. However, overall entropy for all solutions was low (<0.55) indicating inferred clusters were not sufficiently separated, so we did not use these cluster assignments for final analyses.
4) Stability assessment: we evaluated how consistent clusters were when subsampling data, preferring solutions with cluster sizes that were less sensitive to changes in the input data, as our case patients come from voluntary surveillance and thus are likely a sample of all MIS-C cases in the U.S. We randomly subsampled n=2000 cases from the full dataset 100 times and ran LCA varying the number of clusters from 2 to 6. For each cluster number, we used the subsampled LCA models to predict class membership for the full dataset and then computed the Adjusted Rand Index, a measure of similarity between two clustering assignments with 1 indicating identical results, for each pair of subsample-derived models. This allowed us to characterize the stability of inferred clusters. We observed that LCA solutions with cluster sizes of two and three produced the most consistent estimates, as measured by higher Rand Index values, when the input data were varied in this manner (Supplementary Figure 3).
5) Clinical interpretability: In consultation with clinical partners, we determined that the solutions with two to four clusters generally corresponded to interpretable and clinically useful categories.

We therefore selected the LCA solution with three clusters for further investigation on the basis of these criteria: diminishing returns in BIC, maximum entropy, results from LCA integrating variable selection, consistency under subsampling, and clinical interpretability.

### Supplementary Tables and Figures

**Supplementary Table 1. Model fit metrics (degrees of freedom, number of parameters, entropy, information metrics, log-likelihood) versus number of clusters.**

| Clusters | Degrees of freedom | Num params | entropy | BIC | ABIC | AIC | CAIC | Log-likelihood |
|---|---|---|---|---|---|---|---|---|
| 1 | 8915 | 29 | | 273092.7 | 273000.6 | 272886.9 | 273121.7 | -136414.4 |
| 2 | 8885 | 59 | 0.6930307 | 265737.0 | 265549.5 | 265318.1 | 265796.0 | -132600.1 |
| 3 | 8855 | 89 | 0.7673455 | 262591.2 | 262308.4 | 261959.4 | 262680.2 | -130890.7 |
| 4 | 8825 | 119 | 0.6989315 | 261248.1 | 260869.9 | 260403.3 | 261367.1 | -130082.7 |
| 5 | 8795 | 149 | 0.6846483 | 260239.2 | 259765.7 | 259181.5 | 260388.2 | -129441.8 |
| 6 | 8765 | 179 | 0.6781580 | 259566.6 | 258997.8 | 258295.9 | 259745.6 | -128969.0 |
| 7 | 8735 | 209 | 0.6713617 | 259120.8 | 258456.6 | 257637.1 | 259329.8 | -128609.6 |
| 8 | 8705 | 239 | 0.6625687 | 258826.2 | 258066.7 | 257129.6 | 259065.2 | -128325.8 |
| 9 | 8675 | 269 | 0.6671141 | 258652.4 | 257797.6 | 256742.8 | 258921.4 | -128102.4 |
| 10 | 8645 | 299 | 0.6622905 | 258509.1 | 257558.9 | 256386.6 | 258808.1 | -127894.3 |

Vuong-Lo-Mendell-Rubin test *P*-values were <0.0001 for all clusters assessed.

Abbreviations: BIC = Bayesian Information Criterion, ABIC = Sample-size Adjusted Bayesian Information Criterion, AIC = Akaike Information Criterion, CAIC = Consistent Akaike Information Criterion.

**Supplementary Table 2. Distribution of demographics, underlying health conditions, clinical signs, symptoms, complications and laboratory testing results, and clinical outcomes by latent class analysis-inferred clusters.**

| | 1. Respiratory (N=713) | 2. Shock/Cardiac (N=3359) | 3. Undifferentiated (N=4872) | Chi-squared test statistic[a] | P-value[a] |
|---|---|---|---|---|---|
| **Age** | | | | | <0.0001 |
| Median (IQR) | 13 (6.3, 17) | 11 (7.7, 14) | 6.8 (3.6, 10) | | |
| **Age group** | | | | 1400 | <0.0001 |
| <1 y | 45 (7.0%) | 16 (0.5%) | 200 (4.5%) | | |
| 1–4 y | 96 (14.8%) | 283 (9.0%) | 1458 (32.5%) | | |
| 5–9 y | 99 (15.3%) | 1083 (34.3%) | 1627 (36.3%) | | |
| 10–14 y | 162 (25.0%) | 1207 (38.2%) | 905 (20.2%) | | |
| 15–20 y | 245 (37.9%) | 569 (18.0%) | 292 (6.5%) | | |
| **Sex** | | | | 7 | 0.03 |
| Male | 413 (57.9%) | 2086 (62.1%) | 2908 (59.7%) | | |
| **Race/ethnicity** | | | | 137 | <0.0001 |
| Non-Hispanic Asian | 22 (3.2%) | 83 (2.6%) | 143 (3.1%) | | |
| Non-Hispanic Black | 201 (29.0%) | 1194 (37.4%) | 1181 (25.6%) | | |
| Non-Hispanic White | 241 (34.7%) | 1019 (31.9%) | 1823 (39.5%) | | |
| Other/Multiple race | 21 (3.0%) | 139 (4.4%) | 227 (4.9%) | | |
| **Preceding COVID-19-like illness** | 426 (68.5%) | 1413 (49.8%) | 2052 (50.1%) | 78 | <0.0001 |
| **Any pre-existing conditions** | 293 (41.2%) | 1049 (31.4%) | 868 (18.0%) | 301 | <0.0001 |
| Obesity | 181 (25.8%) | 683 (20.6%) | 423 (8.8%) | 295 | <0.0001 |
| Chronic lung disease | 67 (9.6%) | 274 (8.3%) | 236 (4.9%) | 48 | <0.0001 |
| Other congenital malformations | 69 (10.0%) | 122 (3.7%) | 171 (3.6%) | 65 | <0.0001 |
| Seizures | 35 (5.1%) | 65 (2.0%) | 76 (1.6%) | 37 | <0.0001 |
| Congenital heart disease | 23 (3.3%) | 76 (2.3%) | 52 (1.1%) | 29 | <0.0001 |

| | 1. Respiratory (N=713) | 2. Shock/Cardiac (N=3359) | 3. Undifferentiated (N=4872) | Chi-squared test statistic[a] | P-value[a] |
|---|---|---|---|---|---|
| Immunosuppression | 23 (3.3%) | 23 (0.7%) | 30 (0.6%) | 53 | <0.0001 |
| Diabetes mellitus type 1 or 2 | 11 (1.5%) | 37 (1.1%) | 16 (0.3%) | 24 | <0.0001 |
| Sickle cell disease | 14 (2.0%) | 18 (0.6%) | 20 (0.4%) | 27 | <0.0001 |
| **Number of organ systems involved** | | | | | <0.0001 |
| Median (IQR) | 3.0 (3.0, 4.0) | 5.0 (4.0, 5.0) | 4.0 (3.0, 4.0) | | |
| **Clinical signs, symptoms, complications, and laboratory testing results** | | | | | |
| Arrythmia | 148 (21.1%) | 1055 (31.8%) | 788 (16.4%) | 269 | <0.0001 |
| Myocarditis | 23 (3.3%) | 936 (28.3%) | 173 (3.6%) | 1137 | <0.0001 |
| Shock | 59 (8.3%) | 2310 (69.0%) | 479 (9.8%) | 3389 | <0.0001 |
| Elevated troponin | 109 (16.2%) | 2796 (85.7%) | 1488 (31.9%) | 2580 | <0.0001 |
| Elevated BNP or NT-proBNP | 215 (34.6%) | 2694 (94.9%) | 2946 (72.2%) | 1217 | <0.0001 |
| Acute kidney injury | 82 (11.5%) | 1389 (41.7%) | 193 (4.0%) | 1861 | <0.0001 |
| Cough | 582 (82.7%) | 1192 (35.8%) | 1292 (26.6%) | 859 | <0.0001 |
| Shortness of breath | 523 (74.9%) | 1523 (45.6%) | 326 (6.7%) | 2426 | <0.0001 |
| Chest pain/tightness | 280 (40.6%) | 840 (25.3%) | 224 (4.6%) | 1026 | <0.0001 |
| Pneumonia | 465 (65.3%) | 734 (21.9%) | 186 (3.8%) | 1960 | <0.0001 |
| ARDS | 140 (19.7%) | 325 (9.7%) | 20 (0.4%) | 640 | <0.0001 |
| Elevated d-dimer | 551 (80.6%) | 3054 (94.3%) | 4099 (88.4%) | 146 | <0.0001 |
| Thrombophilia | 90 (13.1%) | 534 (16.7%) | 574 (12.5%) | 29 | <0.0001 |
| Thrombocytopenia | 135 (19.3%) | 1837 (55.6%) | 1622 (33.8%) | 530 | <0.0001 |
| Abdominal pain | 290 (42.2%) | 2750 (82.4%) | 2921 (60.5%) | 645 | <0.0001 |
| Vomiting | 395 (55.7%) | 2622 (78.4%) | 2955 (60.9%) | 318 | <0.0001 |
| Diarrhea | 286 (40.3%) | 2240 (67.0%) | 2297 (47.4%) | 366 | <0.0001 |

| | 1. Respiratory (N=713) | 2. Shock/Cardiac (N=3359) | 3. Undifferentiated (N=4872) | Chi-squared test statistic[a] | P-value[a] |
|---|---|---|---|---|---|
| Elevated bilirubin | 102 (14.6%) | 1175 (35.7%) | 641 (13.4%) | 593 | <0.0001 |
| Elevated liver enzymes | 336 (47.9%) | 2290 (69.3%) | 2046 (42.6%) | 569 | <0.0001 |
| Rash | 116 (16.3%) | 1722 (51.4%) | 3143 (64.8%) | 636 | <0.0001 |
| Lesions | 14 (2.0%) | 624 (18.7%) | 1278 (26.5%) | 245 | <0.0001 |
| Headache | 222 (32.2%) | 1925 (57.9%) | 1842 (38.1%) | 362 | <0.0001 |
| Altered mental state | 69 (9.8%) | 669 (20.1%) | 247 (5.1%) | 451 | <0.0001 |
| Syncope/near syncope | 44 (6.3%) | 297 (9.1%) | 103 (2.2%) | 190 | <0.0001 |
| Neck pain | 28 (4.1%) | 1082 (33.3%) | 1002 (21.3%) | 314 | <0.0001 |
| Myalgia | 164 (23.6%) | 1370 (41.3%) | 1103 (23.0%) | 328 | <0.0001 |
| Conjunctival injection | 43 (6.0%) | 1961 (58.6%) | 2874 (59.2%) | 742 | <0.0001 |
| Periorbital edema | 9 (1.3%) | 380 (11.6%) | 668 (14.3%) | 98 | <0.0001 |
| Cervical lymphadenopathy | 22 (3.4%) | 457 (16.0%) | 764 (18.6%) | 97 | <0.0001 |
| **ICU admission** | 353 (49.5%) | 2765 (82.3%) | 1609 (33.0%) | 1942 | <0.0001 |
| **Death** | 33 (4.6%) | 34 (1.0%) | 3 (0.1%) | 170 | <0.0001 |
| **ICU length of stay** | | | | | <0.0001 |
| Median (IQR) | 4.0 (2.0, 7.0) | 4.0 (2.0, 6.0) | 3.0 (1.0, 4.0) | | |
| **Hospital length of stay** | | | | | <0.0001 |
| Median (IQR) | 5.0 (4.0, 8.0) | 7.0 (5.0, 9.0) | 5.0 (3.0, 6.0) | | |

[a] Statistical significance of global differences across clusters was assessed using the Chi-squared test for categorical variables and Kruskal-Wallis test for continuous variables. Chi-squared test statistics were included for categorical variables.

Prevalence was calculated based on non-missing data. Abbreviations: MIS-C = multisystem inflammatory syndrome in children. ICU = intensive care unit. ARDS = acute respiratory distress syndrome. BNP = B-type natriuretic peptide. NT-proBNP = N-terminal prohormone of brain natriuretic peptide.

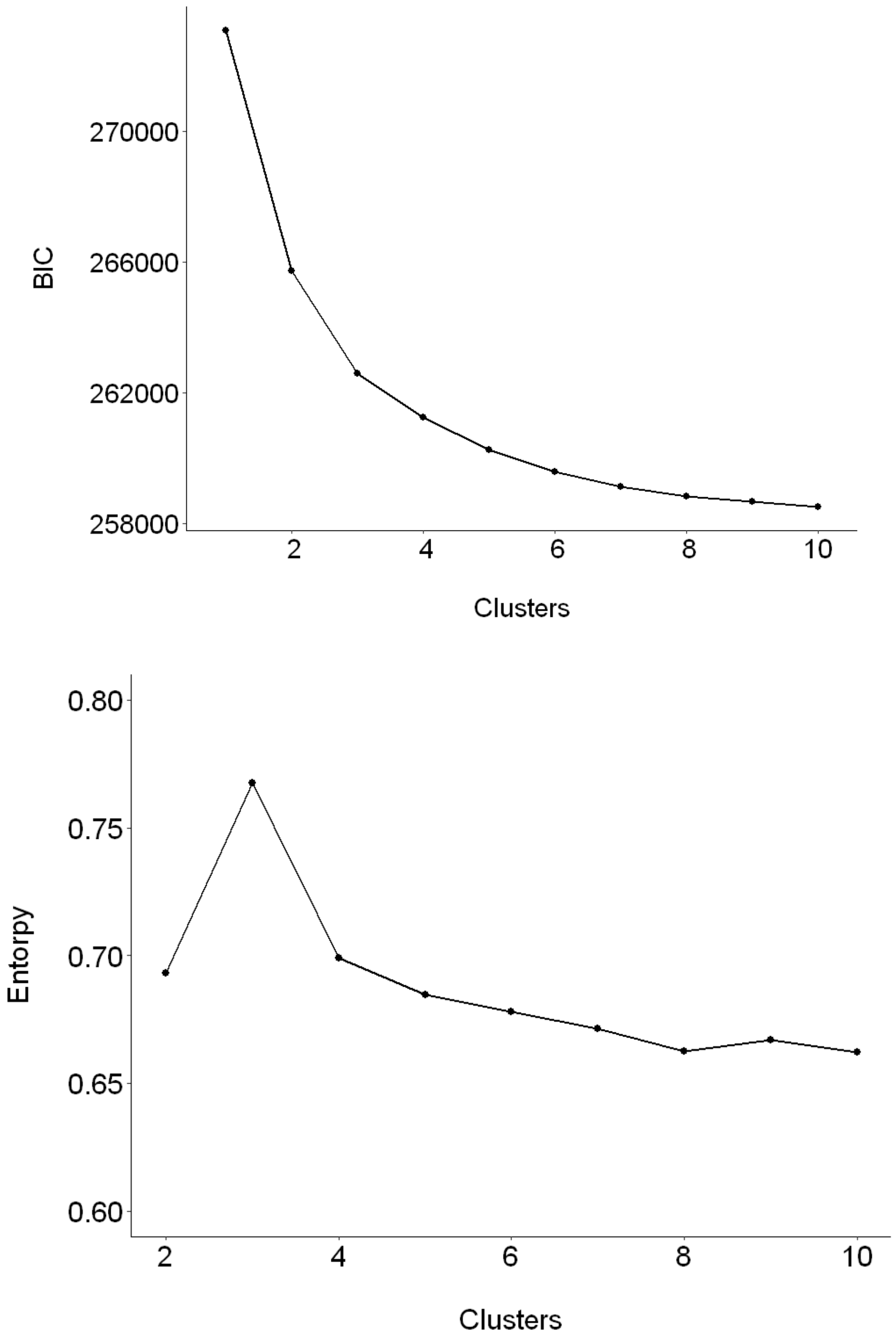

**Supplementary Figure 1. Bayesian Information Criterion (BIC) values (a) and entropy (b) versus number of clusters.** We calculated the Bayesian Information Criterion (BIC) for clusters of size one to ten. BIC continued to decrease for each additional class added at least up until ten clusters. Decreases in BIC began to diminish beginning with clusters of size three to five.

We calculated relative entropy, which is a measure of cluster distinctiveness ranging from 0 to 1 with higher values indicating greater separation between clusters [1], for clusters of size two to ten. An LCA solution with three clusters had the highest entropy (0.77).

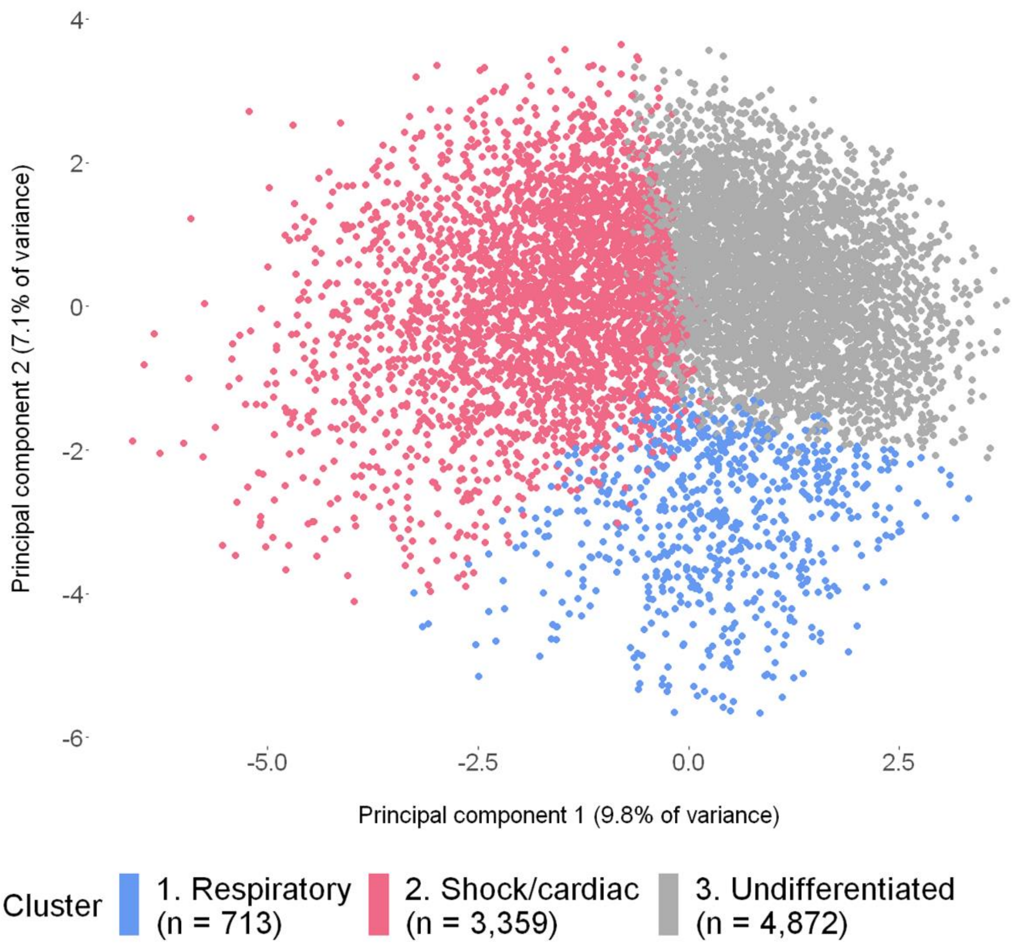

**Supplementary Figure 2. Principal component analysis (PCA) visualization of MIS-C cases by latent class analysis-inferred cluster.** PCA is a variable reduction approach intended to simplify complex datasets into a smaller number of variables (i.e., principal components) that contribute the most variance [2]. PCA indicated visual separation in a plot of MIS-C cases along principal components one (x-axis) and two (y-axis) between the three clusters inferred using LCA. Principal components one and two explain only 17% of the total variance, and therefore are a limited representation of the full variation in MIS-C clinical phenotypes. Principal component one correlated most strongly with shock, elevated troponin, and acute kidney injury, and principal component two correlated most strongly with conjunctival injection, pneumonia, and rash.

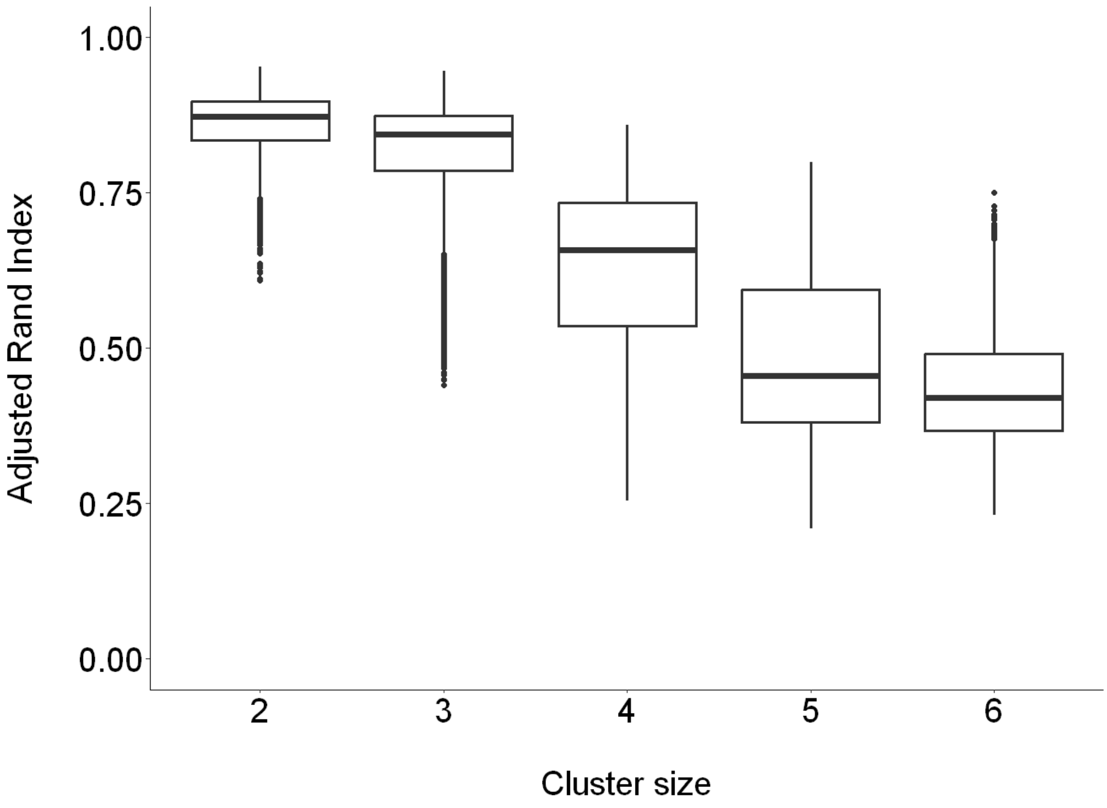

**Supplementary Figure 3. Consistency of latent class analysis-inferred clusters estimated from sub-sampled datasets.** We evaluated how consistent clusters were when subsampling data, preferring solutions with cluster sizes that were less sensitive to changes in the input data, as our case patients come from voluntary surveillance and thus are likely a sample of all MIS-C cases in the U.S. We randomly subsampled n=2000 cases from the dataset 100 times and ran latent class analysis varying the number of clusters from 2 to 6. For each value of cluster size, we used the subsampled LCA models to predict class membership for the full dataset and computed Adjusted Rand Indices, a measure of similarity between two clustering assignments with 1 indicating identical results, for each pair of subsampled models. We observed that LCA solutions with cluster sizes of two and three produced the most consistent estimates, as measured by higher Rand Index values, when the input data were varied in this manner.
