## Supplementary material for "Phenotypic Classification of Multisystem Inflammatory Syndrome in Children: A Latent Class Analysis": Case report form

U.S. DEPARTMENT OF
HEALTH AND HUMAN SERVICES
CENTERS FOR DISEASE CONTROL
AND PREVENTION
ATLANTA, GA 30329

### Multisystem Inflammatory Syndrome Associated with COVID-19 Case Report Form

**MIS ID** *(REQUIRED)*: ______________ **Health Department ID:** ______________

**NNDSS ID** *(local_record_id/case id)*: ______________ **Tools for CRF data submission to supplement NNDSS case notification/data:** ○ DCIPHER ○ RedCap

**Abstractor name:** ______________ **NCOV ID** *(if available)*: ______________ **Date of abstraction:** ______________

#### SECTION 1 – INCLUSION CRITERIA

**1.1** ☐ Age <21, AND

**1.2** ☐ Fever >38.0°C for ≥24 hours, or report of subjective fever lasting ≥24 hours, AND

**1.3** ☐ Laboratory markers of inflammation (including, but not limited to one or more; an elevated C-reactive protein (CRP), erythrocyte sedimentation rate (ESR), fibrinogen, procalcitonin, d-dimer, ferritin, lactic acid dehydrogenase (LDH), or interleukin 6 (IL-6), elevated neutrophils, reduced lymphocytes and low albumin, AND

**1.4** ☐ Evidence of clinically severe illness requiring hospitalization, with multisystem (≥2) organ involvement *(check all applicable below)*: AND
- **1.4.1** ☐ Cardiac (e.g. shock, elevated troponin, BNP, abnormal echocardiogram, arrhythmia)
- **1.4.2** ☐ Renal (e.g. acute kidney injury or renal failure)
- **1.4.3** ☐ Respiratory (e.g. pneumonia, ARDS, pulmonary embolism)
- **1.4.4** ☐ Hematologic (e.g. elevated D-dimers, thrombophilia, or thrombocytopenia)
- **1.4.5** ☐ Gastrointestinal (e.g. elevated bilirubin, elevated liver enzymes, or diarrhea)
- **1.4.6** ☐ Dermatologic, (e.g. rash, mucocutaneous lesions)
- **1.4.7** ☐ Neurological, (e.g. CVA, aseptic meningitis, encephalopathy)

**1.5** ☐ No alternative plausible diagnosis; AND

**1.6** ☐ Positive for current or recent SARS-COV-2 infection by (check all applicable below): OR
- **1.6.1** ☐ RT-PCR
- **1.6.2** ☐ Serology
- **1.6.3** ☐ Antigen test

**1.7** ☐ COVID-19 exposure within the 4 weeks prior to the onset of symptoms
- **1.7.1** If yes, date of first exposure within the 4 weeks prior : *(MM/DD/YYYY)*: ______________ ☐ Unknown

#### SECTION 2 – PATIENT DEMOGRAPHICS

**2.1** **State of Residence:** ______________

**2.2** **Patient zip code**/postal code *(primary residence)*: __________

**2.3** **Date of birth** *(MM/DD/YYYY)*: ______________

**2.4** **Sex:** ○ Male ○ Female

**2.5** **Ethnicity:** ○ Hispanic or Latino ○ Not Hispanic or Latino ○ Refused or Unknown

**2.6** **Race** *(mark all that apply, selecting more than one option as necessary)*:
- **2.6.1** ☐ White
- **2.6.2** ☐ Black or African American
- **2.6.3** ☐ American Indian
- **2.6.4** ☐ Alaska Native or Aboriginal Canadian
- **2.6.5** ☐ Native Hawaiian
- **2.6.6** ☐ Other Pacific Islander
- **2.6.7** ☐ Asian
- **2.6.8** ☐ Other
- **2.6.9** ☐ Refused or Don't know

**2.7** **Height:** __________ inches

**2.8** **Weight:** __________ lbs

**2.9** **BMI:** __________

***Comorbidities:***

| | | | |
|---|---|---|---|
| **2.10.1** | Immunosuppressive disorder/malignancy | ○ Yes | ○ No |
| **2.10.2** | Obesity | ○ Yes | ○ No |
| **2. 10.3** | Type 1 diabetes | ○ Yes | ○ No |
| **2. 10.4** | Type 2 diabetes | ○ Yes | ○ No |
| **2. 10.5** | Seizures | ○ Yes | ○ No |
| **2. 10.6** | Congenital heart disease | ○ Yes | ○ No |
| **2. 10.7** | Sickle cell disease | ○ Yes | ○ No |
| **2. 10.8** | Chronic lung disease | ○ Yes | ○ No |
| **2. 10.9** | Other congenital malformations | ○ Yes | ○ No |
| **2. 10.10** | Other *(specify)*: ______________ | | |

**2.11** Hospital admission date *(MM/DD/YYYY)*: ______________
- **2.11.1** Number of days in the hospital: __________

**2.12** If admitted to the ICU, admission date *(MM/DD/YYYY)*: ______________
- **2.12.1** Number of days in the ICU: __________

**2.13** **Patient outcome:** ○ Died ○ Discharged ○ Still admitted
- **2.13.2** Hospital discharge or death date *(MM/DD/YYYY)*: ______________

#### SECTION 3 – CLINICAL SIGNS AND SYMPTOMS

**3.1** Did the patient have preceding COVID-like illness? ○ Yes ○ No

**3.1.1** Date of symptom onset *(MM/DD/YYYY)*: ______________

**3.2** Date of symptom onset of MIS *(MM/DD/YYYY)*: ______________

**3.3.** Fever ≥ 38.0°C: ○ Yes ○ No

**3.3.1** Date of fever onset *(MM/DD/YYYY)*: ______________

**3.3.2** Highest Temperature: _____ °C

**3.3.3** Number of days febrile: _____

##### Signs and symptoms ***during present illness***

**3.4.1 Cardiac**

| | | | |
|---|---|---|---|
| **3.4.1.1** | Shock | ○ Yes | ○ No |
| **3.4.1.2** | Elevated troponin | ○ Yes | ○ No |
| **3.4.1.3** | Elevated BNP or NT-proBNP | ○ Yes | ○ No |

**3.4.2 Renal**

| | | | |
|---|---|---|---|
| **3.4.2.1** | Acute kidney injury | ○ Yes | ○ No |
| **3.4.2.2** | Renal failure | ○ Yes | ○ No |

**3.4.3 Respiratory**

| | | | |
|---|---|---|---|
| **3.4.3.1** | Cough | ○ Yes | ○ No |
| **3.4.3.2** | Shortness of breath | ○ Yes | ○ No |
| **3.4.3.3** | Chest pain/tightness | ○ Yes | ○ No |
| **3.4.3.4** | Pneumonia | ○ Yes | ○ No |
| **3.4.3.5** | ARDS | ○ Yes | ○ No |
| **3.4.3.6** | Pulmonary embolism | ○ Yes | ○ No |

**3.4.4 Hematologic**

| | | | |
|---|---|---|---|
| **3.4.4.1** | Elevated D-dimers | ○ Yes | ○ No |
| **3.4.4.2** | Thrombophilia | ○ Yes | ○ No |
| **3.4.4.3** | Thrombocytopenia | ○ Yes | ○ No |

**3.4.5 Gastrointestinal**

| | | | |
|---|---|---|---|
| **3.4.5.1** | Abdominal pain | ○ Yes | ○ No |
| **3.4.5.2** | Vomiting | ○ Yes | ○ No |
| **3.4.5.3** | Diarrhea | ○ Yes | ○ No |
| **3.4.5.4** | Elevated bilirubin | ○ Yes | ○ No |
| **3.4.5.5** | Elevated liver enzymes | ○ Yes | ○ No |

**3.4.6 Dermatologic**

| | | | |
|---|---|---|---|
| **3.4.6.1** | Rash | ○ Yes | ○ No |
| **3.4.6.2** | Mucocutaneous lesions | ○ Yes | ○ No |

**3.4.7 Neurological**

| | | | |
|---|---|---|---|
| **3.4.7.1** | Headache | ○ Yes | ○ No |
| **3.4.7.2** | Altered mental state | ○ Yes | ○ No |
| **3.4.7.3** | Syncope/near syncope | ○ Yes | ○ No |
| **3.4.7.5** | Meningitis | ○ Yes | ○ No |
| **3.4.7.6** | Encephalopathy | ○ Yes | ○ No |

**3.4.8 Other**

| | | | |
|---|---|---|---|
| **3.4.8.1** | Neck pain | ○ Yes | ○ No |
| **3.4.8.2** | Myalgia | ○ Yes | ○ No |
| **3.4.8.3** | Conjunctival injection | ○ Yes | ○ No |
| **3.4.8.4** | Periorbital edema | ○ Yes | ○ No |
| **3.4.8.5** | Cervical lymphadenopathy >1.5 cm diameter | ○ Yes | ○ No |

#### SECTION 4 – COMPLICATIONS

| | | | |
|---|---|---|---|
| **4.1** | **Arrhythmia** | ○ Yes | ○ No |
| | If yes: | | |
| **4.1.1** | Ventricular arrhythmia: | ○ Yes | ○ No |
| **4.1.2** | Supraventricular arrhythmia: | ○ Yes | ○ No |
| **4.1.3** | Other arrhythmia *(specify)*: ______________________ | ○ Yes | ○ No |
| **4.2** | Congestive heart failure | ○ Yes | ○ No |
| **4.3** | Myocarditis | ○ Yes | ○ No |
| **4.4** | Pericarditis | ○ Yes | ○ No |
| **4.5** | Liver failure | ○ Yes | ○ No |
| **4.6** | Deep vein thrombosis or PE | ○ Yes | ○ No |
| **4.7** | ARDS | ○ Yes | ○ No |
| **4.8** | Pneumonia | ○ Yes | ○ No |
| **4.9** | CVA or stroke | ○ Yes | ○ No |
| **4.10** | Encephalitis or aseptic meningitis | ○ Yes | ○ No |
| **4.11** | Shock | ○ Yes | ○ No |
| **4.12** | Hypotension | ○ Yes | ○ No |

#### SECTION 5 – TREATMENTS

| | | | |
|---|---|---|---|
| **5.1** | Low flow nasal cannula | ○ Yes | ○ No |
| **5.2** | High flow nasal cannula | ○ Yes | ○ No |
| **5.3** | Non-invasive ventilation | ○ Yes | ○ No |
| **5.4** | Intubation | ○ Yes | ○ No |
| **5.5** | Mechanical ventilation | ○ Yes | ○ No |
| **5.6** | ECMO | ○ Yes | ○ No |
| **5.7** | Vasoactive medications (e.g. epinephrine, milrinone, norepinephrine, or vasopressin) *(specify)*: ______________________ | ○ Yes | ○ No |
| **5.8** | Steroids | ○ Yes | ○ No |
| **5.9** | Immune modulators (e.g. anakinra, tocilizumab) *(specify)*: ______________________ | ○ Yes | ○ No |
| **5.10** | Antiplatelets (e.g. aspirin, clopidogrel) *(specify)*: ______________________ | ○ Yes | ○ No |
| **5.11** | Anticoagulation (e.g. heparin, enoxaparin, warfarin) *(specify)*: ______________________ | ○ Yes | ○ No |
| **5.12** | Dialysis | ○ Yes | ○ No |
| **5.13** | First IVIG | ○ Yes | ○ No |
| **5.14** | Second IVIG | ○ Yes | ○ No |

#### SECTION 6 – STUDIES

**6.1 Blood Test Results**

| | | | | | | |
|---|---|---|---|---|---|---|
| **6.1.1** | Fibrinogen | Highest value: ________ | units: ________ | ○ Low | ○ Normal | ○ High |
| **6.1.2** | CRP | Highest value: ________ | units: ________ | ○ Low | ○ Normal | ○ High |
| **6.1.3** | Ferritin | Highest value: ________ | units: ________ | ○ Low | ○ Normal | ○ High |
| **6.1.4** | Troponin | Highest value: ________ | units: ________ | ○ Low | ○ Normal | ○ High |
| **6.1.5** | BNP | Highest value: ________ | units: ________ | ○ Low | ○ Normal | ○ High |
| **6.1.6** | NT-proBNP | Highest value: ________ | units: ________ | ○ Low | ○ Normal | ○ High |
| **6.1.7** | D-dimer | Highest value: ________ | units: ________ | ○ Low | ○ Normal | ○ High |
| **6.1.8** | IL-6 | Highest value: ________ | units: ________ | ○ Low | ○ Normal | ○ High |

| | | | | |
|---|---|---|---|---|
| **6.1.9** | Serum White blood count | Highest value: ________ | Lowest value: ________ | units: ________ |
| **6.1.10** | Platelets | Highest value: ________ | Lowest value: ________ | units: ________ |
| **6.1.11** | Neutrophils | Highest value: ________ | Lowest value: ________ | units: ________ |
| **6.1.12** | Lymphocytes | Highest value: ________ | Lowest value: ________ | units: ________ |
| **6.1.13** | Bands | Highest value: ________ | Lowest value: ________ | units: ________ |

**6.2 CSF Studies**

| | | | | |
|---|---|---|---|---|
| **6.2.1** | White blood count | Highest value: ________ | Lowest value: ________ | units: ________ |
| **6.2.2** | Protein | Highest value: ________ | Lowest value: ________ | units: ________ |
| **6.2.3** | Glucose | Highest value: ________ | Lowest value: ________ | units: ________ |

**6.3 Urinalysis**

| | | | | |
|---|---|---|---|---|
| **6.3.1** | Urine White blood count | Highest value: ________ | Lowest value: ________ | units: ________ |

**6.4 Echocardiogram** (check if seen on ANY echocardiogram)

- **6.4.1** ☐ Not done
- **6.4.2** ☐ Normal results
- **6.4.3** ☐ Coronary artery aneurysms
  - **6.4.3.1** Max coronary artery Z-score: ________
- **6.4.4** ☐ Coronary artery dilatation
- **6.4.5** ☐ Cardiac dysfunction (decreased function), specify type:
  - **6.4.5.1** ☐ left ventricular dysfunction
  - **6.4.5.2** ☐ right ventricular dysfunction
- **6.4.6** ☐ Pericardial effusion
- **6.4.7** ☐ Pleural effusion
- **6.4.8** ☐ Mitral regurgitation, specify type: ○ mild ○ moderate ○ severe
- **6.4.9** ☐ Other *(specify)*: ________________

**6.5 Date of first test showing coronary artery aneurysm or dilatation** *(MM/DD/YYYY)*: ____________

**6.6 Abdominal imaging** ☐ Ultrasound ☐ CT ☐ Not done

- **6.6.1** ☐ Normal
- **6.6.2** ☐ Mesenteric lymphadenopathy
- **6.6.3** ☐ Free fluid
- **6.6.4** ☐ Other *(specify)*: ________________

**6.7 Chest imaging** ☐ Chest x-ray ☐ CT ☐ Not done

- **6.7.1** ☐ Normal
- **6.7.2** ☐ Pneumonia
- **6.7.3** ☐ Atelectasis
- **6.7.4** ☐ Pleural effusion
- **6.7.5** ☐ Other *(specify)*: ________________

##### SARS-COV-2 testing

**6.8 RT-PCR:** ○ Positive ○ Negative ○ Not done
**6.8.1** If performed, date *(MM/DD/YYYY)*: ____________

**6.9 Antigen:** ○ Positive ○ Negative ○ Not done
**6.9.1** If performed, date *(MM/DD/YYYY)*: ____________

**6.10 IgG:** ○ Positive ○ Negative ○ Not done
**6.10.1** If performed, date *(MM/DD/YYYY)*: ____________

**6.11 IgM:** ○ Positive ○ Negative ○ Not done
**6.11.1** If performed, date *(MM/DD/YYYY)*: ____________

**6.12 IgA:** ○ Positive ○ Negative ○ Not done
**6.12.1** If performed, date *(MM/DD/YYYY)*: ____________

#### SECTION 7 COVID-19 VACCINE INFORMATION

**7.1** **Has the patient received a COVID-19 vaccine?** ○ Yes ○ No ○ Unknown

**7.2** **If yes, how many doses?** ○ 1 dose ○ 2 doses ○ Unknown

**7.2.1** Date dose 1 received *(MM/DD/YYYY)*: ______________

**7.2.2** Date dose 2 received *(MM/DD/YYYY)*: ______________

**7.3** **COVID-19 Vaccine manufacturer** ○ Pfizer ○ Moderna ○ Johnson & Johnson/Janssen

○ Other, *(specify)*: ____________________________________ ○ Unknown

RESET FORM
